# Projected Cost-beneficial Impact of the Selective Cytopheretic Device in Pediatric Acute Kidney Injury Requiring Kidney Replacement Therapy

**DOI:** 10.1101/2025.01.27.25320922

**Authors:** Nathan Kleinman, Jennifer Kammerer, Sai Prasad N. Iyer, Stuart L. Goldstein, Alec Kleinman, Kevin K. Chung, Charuhas V. Thakar

## Abstract

**Rationale & Objective:** The Selective Cytopheretic Device for Pediatrics (SCD-PED) is a cell-directed extracorporeal therapy approved by US FDA for pediatric patients with acute kidney injury (AKI) due to sepsis or a septic condition, requiring antibiotics and continuous renal replacement therapy (CRRT). This study aimed to estimate hospitalization costs and outcomes of SCD-PED therapy by leveraging the Kids’ Inpatient Database (KID) and SCD-PED studies.

**Study Design:** Publicly available hospitalization cost data were combined with clinical metrics from prior SCD-PED studies to assess the impact of SCD-PED on inpatient hospital costs among pediatric patients receiving CRRT.

**Setting & Population:** The SCD-PED was evaluated in two multicenter pediatric studies, involving 16 and 6 patients, respectively. Pediatric patients with AKI and multi-organ dysfunction receiving CRRT as part of standard care were included. The KID subset comprised hospitalizations with CRRT, a length of stay (LOS) up to 60 days, mortality and severity level 4, an AKI diagnosis, and total parenteral nutrition (TPN) procedures.

**Interventions:** Patients received SCD-PED therapy for up to 7 or 10 days or until CRRT termination.

**Outcomes:** Outcomes analyzed included hospital LOS, mortality, vasopressor use, mechanical ventilation, sepsis diagnosis, number of SCD-PED devices used, and hospitalization cost estimates.

**Model, Perspective, & Timeframe:** A regression-based economic cost model compared costs between SCD-PED therapy and theoretical controls, adjusted to 2024 US dollars.

**Results:** Modeled hospitalization costs were $457,092 in the KID cohort and $389,451 in the ppCRRT cohort. Median hospital LOS was lower in the SCD-PED group (28 days vs. 31 days), resulting in lower estimated costs ($320,304) and an estimated savings of $69,146 per hospitalization.

**Limitations:** Small sample sizes and single-arm design with no prospective control arm. Reported costs are estimates based on models.

**Conclusions:** The SCD-PED shows potential for survival benefit and cost-benefit in critically ill children with AKI requiring CRRT, including those with sepsis.

## Rationale & Objective

Sepsis is a leading cause of morbidity and mortality in children [1, 2]. An estimated 22 cases of sepsis occur per 100 000 person-years in children globally, accounting for 4% of hospitalized children and 8% of pediatric intensive care unit (ICU) patients [1]. In the US, it is estimated that at least 200 children are diagnosed with severe sepsis daily, amounting to over 75 000 cases annually [3]. The mortality rate among children with sepsis ranges from 4% to 50%, the higher death rates occurring in patients with multi-organ failures [1].

Acute kidney injury (AKI) is common in children with sepsis, occurring in 42% of cases [4]. Similarly, sepsis is associated with approximately 50% of pediatric AKI cases in the ICU [5]. Compared to pediatric patients with no AKI or mild AKI (Stage 1) associated with sepsis, severe AKI (Stage 2/3) that occurs in the setting of sepsis is associated with increased risk of death, longer ICU length of stay (LOS), longer overall hospital LOS, longer duration of mechanical ventilation use, greater likelihood of blood product use, lower likelihood of returning to baseline renal function, and poorer 3-month post-discharge health-related quality of life (HRQL) or mortality in patients surviving beyond 28 days [6]. The contemporary Worldwide Exploration of Renal Replacement Outcomes Collaborative in Kidney Disease (WE-ROCK) registry reported similar outcomes [7], representing a critically ill and resource intensive patient population.

The Selective Cytopheretic Device for Pediatrics (SCD-PED) is a cell directed extracorporeal therapy designed to abate the hyperinflammatory milieu by sequestering and immunomodulating highly activated inflammatory leukocytes, followed by their release back into the circulation under two novel conditions: 1) a unique low-shear stress blood flow microenvironment and 2) a regional citrate-induced, low ionized-calcium environment of <0.4 mM, conducive to leukocyte deactivation. The SCD-PED promotes an immunomodulatory effect by selectively reprogramming neutrophils towards apoptosis and shifting monocytes from a pro-inflammatory to a reparative phenotype, leading to a resolution of systemic inflammation towards immune homeostasis. Since immunomodulated leukocytes are released back into circulation, immunosuppression does not occur [8, 9].

A Humanitarian Use Device approval for the SCD-PED was granted on February 21, 2024 based on evidence of safety and probable benefit [10] under the humanitarian device exemption regulatory pathway [10]. The SCD-PED is indicated for treatment of pediatric patients (weight ≥10kg and age ≤22 years) with AKI due to sepsis or a septic condition on antibiotic therapy and requiring kidney replacement therapy [10]. The SCD-PED has been tested in two prospective multicenter pediatric studies, one with 16 patients (SCD-PED-01, median age 12.1 years) and the other with 6 patients (SCD-PED-02, median age 2.1 years), showing high survival rates, including 72.7% survival to ICU discharge, 77.3% survival to 60 days after SCD-PED initiation, 94.4% survival to ICU discharge or 60 days in the subset of patients without extracorporeal membrane oxygenation (ECMO), and 100% dialysis independence and 87.5% of patients returning within normal estimated glomerular filtration rate (eGFR) (>90 mL/min/1.73 m^2^) among survivors at 60 days post SCD-PED initiation [8]. Further study compared these outcomes to historical matched controls from the ppCRRT registry, excluding patients on ECMO, and found significantly better survival in SCD-PED versus ppCRRT (adjusted odds ratio 13.4; P=0.01). Bayesian analysis indicated a 98% probability of higher survival odds in SCD-PED, with a predicted risk difference of 22.4%. There are no other approved selective therapies in the US for pediatric patients with AKI due to sepsis on CRRT.

Given the encouraging survival rates among critically ill children treated with the SCD-PED, the goal of the current study is to determine its financial impact to a health care system from an inpatient perspective. To achieve this goal, we chose to combine publicly available hospitalization cost data with non-cost clinical metrics derived from prior SCD-PED studies to estimate the effect of the SCD-PED on inpatient hospital costs among a pediatric population receiving CRRT.

## Study Design

As cost information was not available for the hospitalizations in the prior SCD-PED studies, this study uses regression-based economic cost modeling to compare the difference in cost between patients using the SCD-PED and CRRT patients not using the SCD-PED. Hospitalization costs were modeled using data from the 2019 Kid’s Inpatient Database (KID) [11]. The KID provides information on hospital stays for children (under age 21) in the United States. It contains data on pediatric hospital admissions, including demographics, diagnoses, and procedures (from the International Classification of Diseases, 10th Revision, Procedure Coding System ICD-10-PCS). The KID offered a nationally representative sample containing data from roughly 3 million pediatric inpatient hospitalizations across 48 US states plus the District of Columbia.

## Setting & Population

The KID subset used in this study included hospitalizations that underwent CRRT, had an overall LOS up to 60 days, had KID mortality risk level 4 and severity level 4, had an AKI diagnosis, and had a total parenteral nutrition (TPN) procedure (see **Figure 1**). A hospitalization was determined to have had CRRT if it included the ICD-10-PCS procedure code 5A1D90Z (Performance of Urinary Filtration, Continuous, Greater than 18 hours Per Day). See **Table 1** for codes used to define other variables. Prior pediatric SCD studies excluded patients weighing less than 10 kg or who were over 22 years old. Patient weight is not available in the KID, so a proxy criterion was made by excluding hospitalizations of children under one year of age. The KID was also limited to hospitalizations of patients up to only 20 years of age. Hospitalizations with missing cost or length of stay information were excluded.

**Figure 1:**
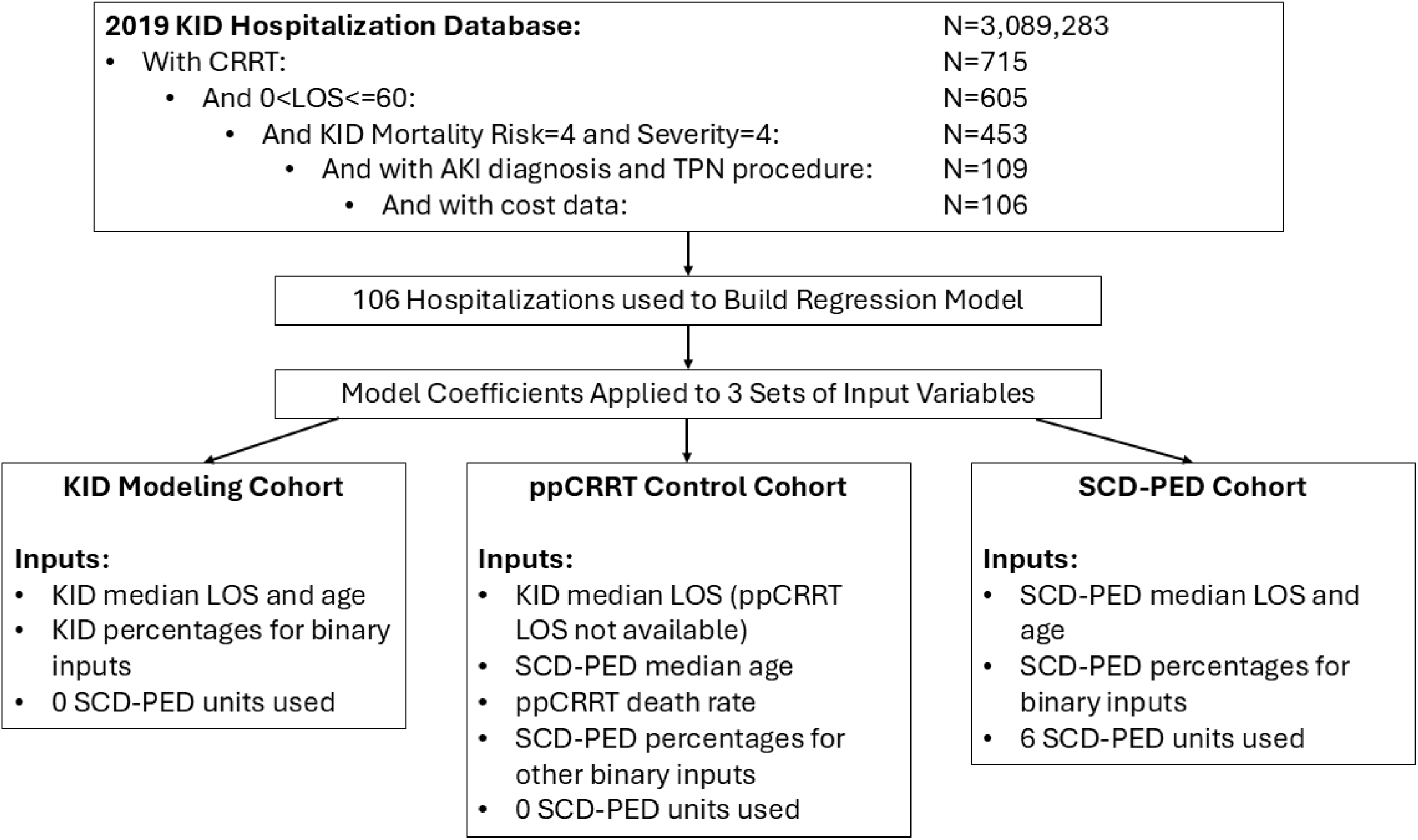
Hospitalization Selection and Model Inputs. KID=Kids Inpatient Database; N=number of hospitalizations; CRRT=continuous renal replacement therapy; LOS=length of stay; AKI=acute kidney injury; TPN=total parenteral nutrition; ppCRRT=Prospective Pediatric CRRT registry; SCD-PED=selective cytopheretic device for pediatrics.

**Table 1:** Descriptive Characteristics of KID Hospitalizations CRRT used in Modeling (N=106)

| **Data Fields** | **Mean** | **Standard Deviation** |
| --- | --- | --- |
| Age (years) (median=12.0) | 11.0 | 6.2 |
| Female | 49.1% | 50.2% |
| Length of Stay (days) (median=31.0) | 30.9 | 14.8 |
| Total Cost (2024 dollars) (median=\$351,719) | \$450 032 | \$334 042 |
| Died During Hospitalization | 42.5% | 49.7% |
| Sepsis (ICD-10 Diagnosis Codes A02.1, A22.7, A26.7, A32.7, A40.0, A40.1, A40.3, A40.8, A40.9, A41, A42.7, A54.86, B00.7, B37.7, O033.7, O038.7, O048.7, O073.7, O08.82, O85, O86.04, O86.81, P36, R65.2) | 75.5% | 43.2% |
| Vasopressor Use (ICD-10 Procedure Codes 3E030XZ, 3E033XZ, 3E040XZ, 3E043XZ, 3E050XZ, 3E053XZ, 3E060XZ, 3E063XZ) | 24.5% | 43.2% |
| Mechanical Ventilation (ICD-10 Diagnosis Codes Z99.11 or J95.85 or ICD-10 Procedure Codes 5A1935Z, 5A1945Z or 5A1955Z) | 81.1% | 39.3% |
| CRRT (ICD-10 Procedure Code 5A1D90Z) | 100.0% | 0.0% |
| AKI (ICD-10 Diagnosis Code N17) | 100.0% | 0.0% |
| Total Parenteral Nutrition (ICD-10 Procedure Codes 3E0406Z, 3E0436Z, 3E0606Z, 3E0636Z) | 100.0% | 0.0% |
| KID Mortality Risk Level 4 | 100.0% | 0.0% |
| KID Severity Level 4 | 100.0% | 0.0% |

CRRT=continuous renal replacement therapy; ICD-10=International Classification of Diseases 10<sup>th</sup> Revision; AKI=acute kidney injury. KID=Kids' Inpatient Database.

The following fields from the KID were used as independent variables in the regression model: hospital LOS, patient age, patient sex, and death indicator. Additionally, the following independent indicator variables were created using ICD-10 procedure and diagnosis codes captured during the KID hospitalization: sepsis diagnosis, vasopressor procedure, and mechanical ventilation procedure. Inpatient cost for each hospitalization was calculated by multiplying the KID’s total charges data field by the KID cost-to-charge ratio.

## Interventions

Patients received SCD-PED therapy for up to 7 days or until CRRT termination in SCD-PED-01, and 10 days or until CRRT termination in SCD-PED-02.

## Outcomes

Outcomes analyzed included hospital LOS, mortality, vasopressor use, mechanical ventilation, sepsis diagnosis, number of SCD-PED devices used, and hospitalization cost estimates.

## Model, Perspective, & Timeframe

A generalized linear regression model with gamma distribution was built in Python^TM^ (Python Software Foundation, Beaverton, Oregon, USA) to model the impact of the independent variables on cost per hospitalization. This type of regression model was chosen for its ability to account for the skewness of cost data. The coefficients from the model were then multiplied by 3 sets of independent variable values (then exponentiating the sum of their products) to obtain cost estimates for 3 hypothetical populations: the KID set of hospitalizations used to build the model, a CRRT non-SCD-PED theoretical control group derived from a matched cohort from the ppCRRT registry, and a hypothetical group of SCD-PED patients (differing from controls in death rate and LOS). The inputs to the model for each population included percentages for the sex, death rate, vasopressor use, mechanical ventilation use, and sepsis variables and median values for age and length of stay (age and length of stay were found to have non-normal distributions using both the Shapiro-Wilk test and the D’Agostino and Pearson’s test). For the SCD-PED population, theoretical device costs were included in the economic model (Figure 1). Hospitalization costs for each population were adjusted to 2024 dollars using the hospital services Consumer Price Index [12].

## Results

The 2019 KID dataset contained 715 hospitalizations that included the CRRT procedure code and where the patient’s age was at least one year. Of these, 598 hospitalizations had LOS<=60. One hundred nine of these had KID mortality risk level 4 and severity level 4, had an AKI diagnosis, and had a TPN procedure. Three of the 109 hospitalizations were missing cost information. Thus, the regression modeling was performed on the remaining 106 hospitalizations. The mean age of the patients from these hospitalizations was 11.0 years (standard deviation [SD] 6.2 years). The average LOS was 30.9 days (SD 14.8), and 42.5% of the patients died before discharge. The mean cost per hospitalization was $450 032 (SD $334 042) in 2024 dollars. Additional descriptive characteristics are provided in **Table 1**.

**Table 2** provides the regression coefficients, standard errors, and p-values for each input variable in the generalized linear model. All input variables were statistically significant (p<0.05) except for age and the indicators for female and sepsis. The variables included in the regression model were used as inputs to the economic model to predict and compare cost per hospitalization for the three hypothetical populations. **Table 3** presents the values used as inputs as well as the resulting cost per hospitalization estimates for each hypothetical population. Most input values for the controls and the SCD-PED hospitalizations were taken from the combined SCD-PED cohort in the studies by Goldstein et al [8]. The 45.2% death rate value for controls was from the ppCRRT non-SCD-PED comparison population used in the Goldstein study. Modeled hospitalization costs were $457 092 in the KID group, reflecting heterogenous, complex cases and costly burden (Table 3). Modeled hospitalization costs were $389 451 in the ppCRRT control group. Median hospital LOS was lower in the SCD-PED group: 28 days vs. 31 days in the ppCRRT group, which when coupled with the lower mortality rate (22.7% vs 45.2%), resulted in a lower estimated cost of $320 304, reflecting an estimated savings of $69 146 per hospitalization. Savings of $39 146 to $46 646 were maintained at theoretical price points of $3750 to $5000 per unit per day for 6 days of therapy (mean duration of therapy from the SCD-PED studies [8]).

**Table 2:** Regression Model Independent Variables and Coefficients.

| Variable | Coefficient | Standard Error | P-Value |
| --- | --- | --- | --- |
| Intercept | 11.153 | 0.276 | <0.001 |
| Length of Stay | 0.034 | 0.005 | <0.001 |
| Age | 0.012 | 0.011 | 0.276 |
| Female | 0.101 | 0.133 | 0.450 |
| Died During Hospitalization | 0.415 | 0.141 | 0.003 |
| Vasopressor Use | -0.475 | 0.159 | 0.003 |
| Mechanical Ventilation | 0.431 | 0.178 | 0.016 |
| Sepsis | -0.086 | 0.155 | 0.578 |

**Table 3:**
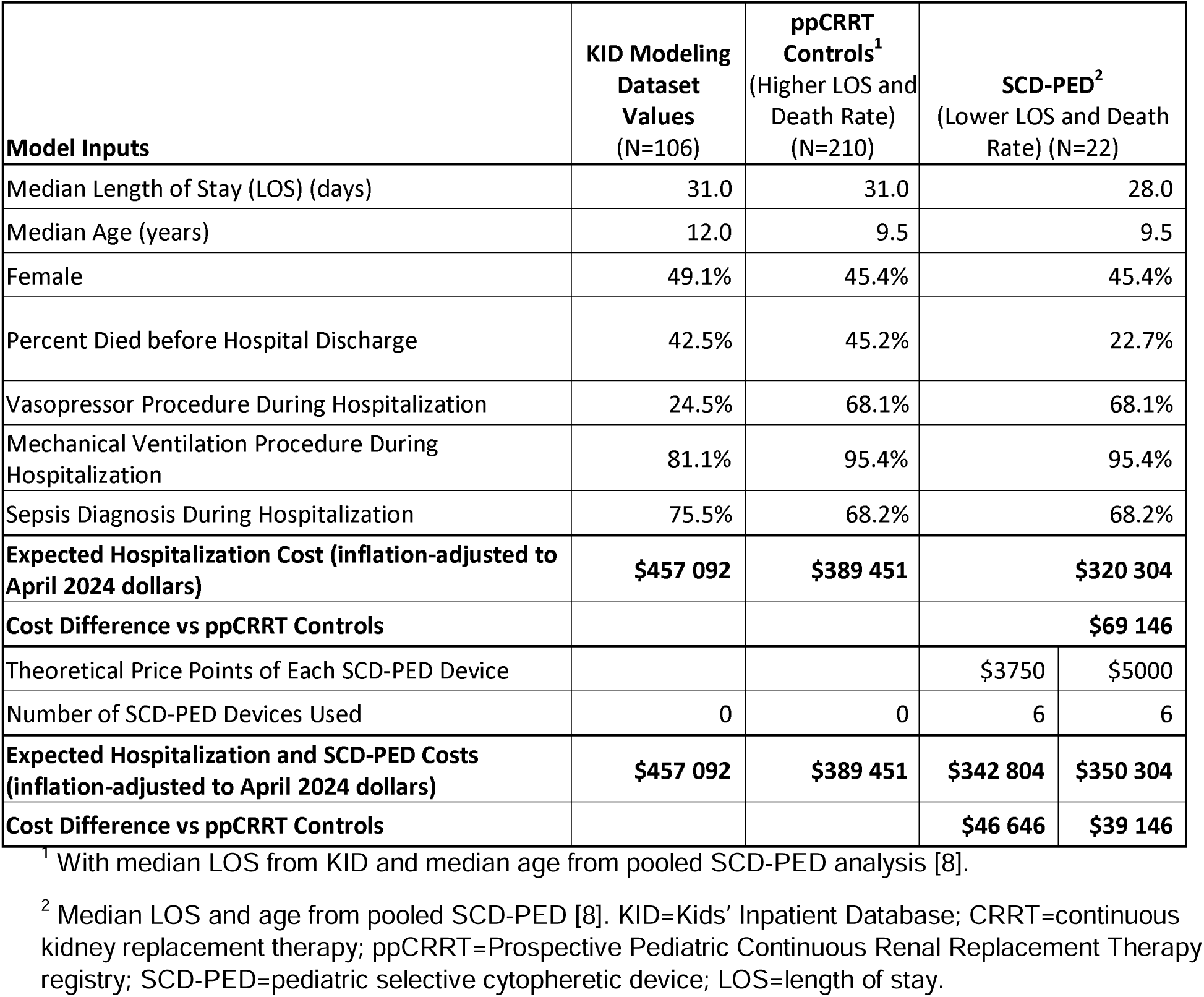
SCD-PED Cost Savings Model in Pediatric AKI Requiring CRRT.

Our analysis found that use of SCD-PED devices would result in both lower death rates and shorter lengths of stay as well as be projected to incur lower costs of hospitalization in critically ill children requiring CRRT. Goldstein et al. found a significant difference in death rate between SCD-PED users (22.7%) and a similar external control group of patients (45.2%, p=0.04) receiving CRRT from the ppCRRT registry [8]. Median LOS from the ppCRRT population was not available, but the 31-day median in the KID model population (used as a proxy for the ppCRRT control cohort LOS based on similar clinical features), was longer than the 28-day median from the SCD-PED study. Using the regression results from the current study, the differences in death rates and LOS correspond to $69 146 lower cost per hospitalization in the SCD-PED cohort. Our estimates demonstrate that with an SCD-PED cost point between $3750 and $5000, there would be projected cost-savings per hospitalization. Potentially, the device could be used for at least 6 days of therapy at cost-neutrality (**Table 3**). Given that the average duration of treatment in the pooled SCD-PED studies [8] was 6, most pediatric patients with AKI requiring CRRT, including those with sepsis, would be treated without imposing any cost to either the institution or the healthcare system, with potential cost-beneficial effects overall.

Given the high death rates found in pediatric patients with AKI and sepsis or sepsis-like conditions and the corresponding high hospitalization costs, the comparative results of Goldstein et al. and the associated cost results from the current study are encouraging and warrant further study.

### Limitations

The current study has several limitations. First, the SCD-PED’s humanitarian approval was for pediatric patients weighing at least 10kg and with age less than or equal to 22 years. The KID data does not contain patient weight, so a surrogate of age≥1 was used. Additionally, the KID’s upper age limit was 20 years. Furthermore, in the initial protocol for the SCD-PED-01 study, patients had to be on CRRT for at least 12 hours to ensure that appropriate ionized calcium levels of <0.4 mM were achieved, and an additional 4 hours before initiation of the SCD, for a total of ∼16 hours of CRRT time [9]. Since we only included patients on CRRT for greater than 18 hours (ICD-PCS procedure code 5A1D90Z) in this analysis, these types of patients would not have been captured by this specific procedure code, but may be through the 5A1D80Z procedure code which would include patients on 6-18 hours of prolonged intermittent hemodialysis. Next, the costs reported herein are estimates based on models; no actual cost information was available from SCD-PED recipients. Finally, the referenced SCD-PED studies had small sample sizes and a single arm design, with no prospective control arm. However, the present analysis provides robust cost estimates by utilizing a nationally representative critically ill pediatric patient sample, and thus offering the results a degree of generalizability.

Further study of the impact of the SCD-PED on outcomes, including mortality, total hospital and ICU length of stay, and cost is warranted. The savings of nearly $70 000 projected with SCD-PED therapy is unlikely to result solely from reduction of 3 days of reductions in LOS. A recent large 2017 analysis of 26 ICUs across 13 hospitals in the US that included over 56 000 critically ill patient records including pediatric (age 17 and over) demonstrated that mean cost of 3.3 days of ICU LOS was $16 353 and the median total cost was $9619 [13]. These costs are likely to be higher when adjusted to current levels; e.g., daily charges of ICU stay at a leading US institution at time of submission (2024) were $11 500 [14]. Interestingly, non-surviving patients who died before discharge had 12.4% higher costs than surviving patients (p<0.01), with a more pronounced effect for patients on MV with longer LOS. The authors postulated that paradoxical findings of survivors costing less than non-surviving patients (“the mortality effect”) was possibly explained by higher resource utilization in patients nearing end of life (e.g. possible increased use of IV vasopressors and blood products etc.). Additionally, terminal patients with do not resuscitate orders may continue to be on MV as a part of comfort care measures, resulting in extended LOS and associated care and resource utilization [13]. Given that nearly every patient in the SCD-PED cohort was on MV (∼95%), the findings from the Kramer study may be relevant to apply here in attempting to explain some the factors that contribute to the projected savings in the SCD-PED cohort. We also speculate that use of SCD-PED, as proposed, may modify the inflammatory parameters and offer different trajectory of organ recovery, thus resulting in overall savings over the course of entire hospitalization.

Furthermore, research on dialysis use and related costs after hospitalization (i.e., readmissions due to sepsis, incident CKD or requirement for long-term dialysis, or other issues) is also needed. For example, there may be significant impact on The Severe Sepsis and Septic Shock Early Management Bundle or SEP-1 and associated quality measures, given its recent inclusion as criteria for reimbursement in the Hospital Value-Based Purchasing (VBP) Program by the Centers for Medicare and Medicaid Services (CMS) [15], and could result in additional cost reductions in overall care. Some of these outcomes (involving up to 90 days of follow-up) will be measured in the prospective QUELIMMUNE (SCD-PED) PediAtric SurVeillance REgistry (SAVE) (NCT06517810), which is currently recruiting.

## Conclusions

In conclusion, the analysis demonstrates likely cost-savings in patients when treated with SCD-PED use in the setting of AKI due to sepsis requiring CRRT. This, along with likely improvements in mortality, demonstrate encouraging implications to both survival and cost-effective advanced therapies in this clinical setting. Future studies should consider direct comparison of both clinical outcomes and economic measures.

## Financial Disclosures

SPI and KKC are employees of SeaStar Medical and receive compensation in the form of salary and company stock. JK is a former employee of SeaStar Medical. NK, JK, SG, AK, and CT receive consulting fees from SeaStar Medical.

## Funding

Funding for this study was provided by SeaStar Medical.

## Data Availability

All data produced in the present study are available upon reasonable request to the authors

## Acknowledgements

Preliminary analysis of the pediatric CRRT economic model was previously presented at American Society of Nephrology’s Kidney Week 2024 Annual Meeting as poster # TH-PO070.

## Authors’ Contributions

All authors made substantial contributions to the design of the study and to the acquisition, analysis, or interpretation of the data. The first draft of the manuscript was prepared by NK and AK. Each author contributed important intellectual content during manuscript drafting or revision and accepts accountability for the overall work by ensuring that questions pertaining to the accuracy or integrity of any portion of the work are appropriately investigated and resolved.

## Data Sharing Statement

All the data in this manuscript is proprietary and owned by SeaStar Medical, Inc. All data are included in the manuscript within the figure and tables.

## Notes

### Clinical Protocols

https://clinicaltrials.gov/study/NCT02820350?intr=Selective%20Cytopheretic%20Device&rank=7

### Author Declarations

IRB of Cincinnati Childrens Hospital Medical Center gave ethical approval for this work. IRB of University of Michigan gave ethical approval for this work. IRB of Childrens Hospital of Alabama gave ethical approval for this work. IRB of Childrens Healthcare of Atlanta gave ethical approval of this work.

